# Genetic liability to insomnia and substance use disorders in patients with bipolar disorder

**DOI:** 10.1101/2024.09.20.24314063

**Authors:** Lindsay M. Melhuish Beaupre, Brandon Coombes, Anthony Batzler, Jorge A. Sanchez-Ruiz, Bhanu Prakash Kolla, Francisco Romo-Nava, Vanessa Pazdernik, Gregory Jenkins, T. Cameron Waller, Michelle Skime, Susan McElroy, Mark Frye, Joanna M. Biernacka

## Abstract

**Background:** Insomnia and substance use disorders (SUD) are common comorbidities of bipolar disorder (BD). Genome-wide association studies (GWAS) have uncovered shared genetic contributions to insomnia and BD as well as SUDs and BD. Using electronic health record (EHR) derived phenotypes (phecodes) and questionnaire data, the authors examined the relationship between insomnia genetic liability and SUDs in BD.

**Methods:** 40,839 participants from the Mayo Clinic Bipolar Disorder Biobank (BD Biobank; n=774) and Mayo Clinic Biobank (n=485 BD cases, n=39,580 controls) were included in the phecode analyses [insomnia, SUD, alcohol use disorder (AUD) and tobacco use disorder (TUD)]. 1789 cases were included in analyses of the BD Biobank questionnaire data, which included information on BD subtype and various SUDs. Logistic regression was used to test for associations between insomnia polygenic risk scores (PRS) and insomnia and SUD phenotypes in BD cases and controls.

**Results:** Insomnia PRS was associated with the insomnia phecode in controls (OR=1.19, p=9.64e-33) but not in BD cases (OR=1, p=0.95). Associations between insomnia PRS and SUD phecodes were significant in BD cases and controls with the effect of the association between insomnia PRS and SUD being stronger in BD cases (interaction p=0.024). In the BD Biobank, the insomnia PRS was associated with increased odds of AUD (OR=1.19, p=4.26e-04), TUD (OR=1.21, p=1.25e-05) and cannabis use disorder (OR=1.16, p=4.19e-03).

**Conclusion:** The effect of genetic predisposition to insomnia on SUD may be stronger in BD cases than controls. This could have clinical treatment implications for individuals with BD and comorbid SUD.

## Introduction

Bipolar disorder (BD) is a chronic psychiatric disorder associated with significant disease burden, driven, in part, by high rates of comorbid medical and psychiatric conditions. Both insomnia and substance use disorders (SUDs) are common comorbidities of BD that frequently co-occur, adversely affect disease course and lead to worse outcomes in those diagnosed with BD, including elevated suicide risk ^1,2^. Thus, it is important to understand the interplay between insomnia and substance use in BD to improve treatment strategies and promote favorable disease outcomes.

Insomnia affects about 40% of individuals with BD ^3^. Insomnia is among the prodromal symptoms of BD and subsequent mood episodes, is associated with more severe episodes, and persists in many individuals during the euthymic period, promoting episode reoccurrence ^4–6^. SUDs are also frequently observed in individuals with BD, with prevalence rates ranging between about 20% for cannabis use disorders (CUD) to higher than 30% for alcohol use disorders (AUD) ^7,8^. Conversely, individuals with a SUD have up to a 13-fold increased risk of developing BD, with risk varying based on the age of onset and severity of the SUD ^9^. Insomnia is also highly prevalent among those with SUDs ^10–13^, with up to 75% of those with alcohol dependence reporting insomnia ^13^. It is a common consequence of substance withdrawal, with one study reporting that 88% of individuals experienced insomnia symptoms during early withdrawal; however, insomnia is also considered a risk factor for the development of a SUD ^11,14^. Yet, the mechanisms underlying the relationship between insomnia and SUDs in patients with BD remains largely unexplained.

Insomnia, BD and SUDs are moderately to highly heritable traits and many genomic loci have been shown to be associated with BD, insomnia and SUDs including AUD, CUD and opioid use disorder (OUD) ^15–19^. Large genome-wide association studies (GWAS) have enabled investigations into the shared genetic architecture across diseases. For example, positive genetic correlations have been shown to exist between insomnia and AUD, insomnia and OUD, as well as BD and insomnia, suggesting shared genetic contributions across diseases ^16–19^. Polygenic risk score (PRS) analyses have also demonstrated that genetic liability to insomnia is associated with BD type II (BD-II) as well as AUD ^20,21^. Together, these studies highlight shared common genetic variation among insomnia and BD as well as insomnia and AUD, motivating us to explore the relationship between insomnia genetic liability and SUDs in BD.

Using data from electronic health records (EHR), we examined associations between insomnia genetic liability and insomnia, SUD, AUD and tobacco use disorder (TUD) in patients with BD and in those without BD. We also analyzed Mayo Clinic Bipolar Disorder Biobank data to examine associations between insomnia genetic liability and specific SUDs in patients with BD.

## Methods and Materials

### Sample

#### Mayo Clinic Biobank (MCB)

The MCB is a cohort of about 58,000 participants recruited at Mayo Clinic who provided a blood sample, completed a questionnaire at enrollment, and consented to use of the collected data along with information from their EHR for research ^22^.

#### Mayo Clinic Bipolar Disorder Biobank (BD Biobank)

Details of the study protocol have been described previously ^23^. The BD Biobank enrolled n=2286 participants with a BD diagnosis ascertained by the clinician rated Structured Clinical Interview for the DSM-IV-TR ^24^. Recruitment sites included Mayo Clinic, Lindner Center of Hope/University of Cincinnati, University of Minnesota, University Hospital of the Universidad Autónoma de Nuevo León (México) and Universidad de Los Andes (Chile). Patient and clinical questionnaires were completed to collect detailed information on demographics, BD subphenotypes, comorbidities and treatment outcomes. Patients also provided a blood sample and consented to use of their EHR data for research, where available.

All MCB and BD Biobank participants provided informed consent for use of their data for research. The Institutional Review Board of Mayo Clinic approved both biobanks and this project.

### Phenotypes of interest

Phenotypes were derived from diagnostic codes in EHR and data collected at time of enrollment into the BD Biobank. Briefly, for EHR-derived phenotypes, ICD-9 and ICD-10-CM (International Classification of Diseases, versions 9 and 10 with clinical modifications) diagnostic codes between January 1^st^ 1985 and April 6^th^ 2020 were mapped to phecodes ^25^, which are clinically meaningful, manually curated and validated, groups of diagnoses ^26,27^. BD cases included clinician-confirmed cases from the BD Biobank and MCB participants having two or more occurrences of phecode 296.1 (BD) in their EHR. Controls were defined as MCB participants with zero occurrences of phecodes 296.1 (BD), 295 (schizophrenic disorders) or 297.1 (schizophrenia). MCB participants with exactly one occurrence of phecode 296.1 were removed from the analysis.

Insomnia and SUDs were identified in EHRs using the following phecodes: 327.4 (insomnia), 316 (substance use and addictions, referred to as substance use disorders), 317.1 (alcoholism, referred to as alcohol use disorders) and 318 (tobacco use disorder). Binary insomnia/substance-related phenotypes were defined based on having at least two occurrences of the corresponding phecode versus zero occurrences. Participants with only one occurrence of the phecodes of interest were removed from relevant analyses. To reduce potential confounding by the amount of available data in the EHR, we limited phecode analyses to those with a record length of at least one year, meaning that the patient had at least two Mayo Clinic visits spaced at least one year apart. Figure S1 visualizes the inclusion process for the combined MCB and BD Biobank EHR-based analysis.

SUD phenotypes were also defined using BD Biobank clinical questionnaire data on whether an individual had a past and/or present SUD at the time of study enrollment. SUDs included in the analyses were: AUD, TUD, CUD, OUD (which combined prescription opioid, other narcotic and heroin use disorders), cocaine use disorder, methamphetamine use disorder and benzodiazepine use disorder. For analyses stratified by BD type I (BD-I) vs. BD-II subtype, we used SCID defined diagnoses assessed at time of BD Biobank enrollment, with schizoaffective disorder BD type (n=40) grouped with BD-I (n=1183).

### Genotyping and Imputations

All MCB and BD Biobank participants recruited at Mayo Clinic were genotyped at the Regeneron Genetics Center (Tarrytown, NY, USA) using Genotyping-by-Sequencing (GxS) technology, which captures “backbone” regions of the genome at lower depths than exome regions. The “backbone” regions then undergo post-processing to enhance genotype quality. The full process is described in Coombes *et al.* 2023 ^28^. Prior genotyping of the BD Biobank was performed in two batches using the Illumina HumanOmniExpress and Global Screening Array, respectively, and included participants recruited outside of the Mayo Clinic site.

Genetic Quality Control (QC) was performed for each genotyping batch separately. Samples were removed for sex discrepancies, heterozygosity <70% on multiple chromosomes, or genotype missingness >5%. Variants were removed if they had a minor allele frequency (MAF) <5%, excessive SNP missingness (>1%) or were not in Hardy-Weinberg Equilibrium (p<1e-6).

Imputation was performed for each batch using the TOPMed imputation server ^29^. Briefly, this server performs haplotype phasing using Eagle2 ^30^ followed by imputation with minimac4 ^31^. We used the TOPMed reference panel that includes n>97,000 sequenced genomes from diverse ancestries ^32^. After imputation, the batches of genotype data were merged into one dataset, keeping only variants with dosage-R^2^ > 0.3. For BD Biobank participants from Mayo Clinic with both GxS and array-based data, the GxS data was used for this study. After merging all the genetic data, relatedness was assessed using KING software ^33^ and one individual was removed from pairs with an estimated second or higher degree of relatedness (kinship coefficient ≥ 0.0442), preferentially keeping individuals with BD over controls; if both subjects had BD then those in the BD Biobank were preferentially kept and if both individuals were controls, then the individual with the longer EHR was kept. Ancestry was estimated using the ADMIXTURE software ^34^ with the 1000 Genomes reference dataset ^35^. Only individuals with >80% proportion European ancestry were kept in the analysis because the PRS used (detailed below) is based on a GWAS including only individuals of European ancestries. FlashPCA ^36^ was used to calculate principal components for each subject.

After QC, relatedness and ancestry removals, as well as restricting to individuals recruited at Mayo Clinic sites, our analyses of EHR-derived phenotypes included 774 participants from the BD Biobank and 40,065 MCB participants (n=485 with BD and n=39,580 controls); the analyses of phenotypes from the BD Biobank questionnaires included 1789 participants with BD enrolled at US sites.

### Polygenic Risk Score Calculations

The merged genetic dataset was restricted to variants that overlapped with HapMap3+ SNPs with MAF >5% and dosage-R^2^ > 0.8 ^37^. Using summary statistics for insomnia from a UK Biobank GWAS ^15^, the insomnia PRS was calculated using LDPred2-auto (v. 1.12.1) ^37^ implemented in the bigsnpr R package, which calculates the optimal sparsity *p* value and SNP heritability. The PRS was standardized to a mean of 0 and standard deviation (SD) of 1.

### Statistical Analyses

Demographic and clinical characteristics were compared between BD cases from the BD Biobank, BD cases from the MCB, and controls from the MCB using analysis of variance for continuous variables and Chi-square tests for categorical variables.

We fit regression models to test for associations between insomnia PRS and dichotomous insomnia and SUD phenotypes in BD cases and controls separately. Sex and the first 10 principal components were included as covariates. We also fit models that included an interaction term between the insomnia PRS and BD case status to test whether associations of the insomnia PRS with insomnia/SUD diagnoses differed between patients with BD and controls.

For the case-only analyses of outcomes derived from BD Biobank questionnaire data, we used regression models to test for associations between insomnia PRS and SUDs as well as BD subtype (BD-I vs. BD-II), while accounting for sex, the top 5 principal components and genotyping batch as covariates. In exploratory analyses, we also tested for association between SUDs and an interaction term between the insomnia PRS and BD subtype to evaluate whether any associations between insomnia PRS and SUD depended on BD subtype. To account for multiple testing, we used a threshold of p<0.01 to determine statistical significance because of the high correlation among phenotypes.

## Results

The sample used for the EHR-derived phenotype analyses was comprised of 40,839 participants of European ancestry with sufficient EHR data (Table 1). Of these, n=774 were cases from the BD Biobank, n=485 were MCB participants diagnosed with BD and n=39,580 were MCB participants without a diagnosis of BD or schizophrenia. BD cases from both the BD Biobank and MCB had significantly higher occurrences of insomnia and SUD phecodes (p<0.001).

**Table 1:** Demographics and phenotype distribution for the combined MCB and BD Biobank sample used in the EHR data analysis.

|  | Mayo Clinic<br>Bipolar<br>Disorder<br>Biobank†<br>(N=774) | Mayo Clinic<br>Biobank -<br>Bipolar<br>Disorder cases<br>(N=485) | Mayo Clinic<br>Biobank<br>(N=39,580) | p value |
| --- | --- | --- | --- | --- |
| **Sex** |  |  |  | < 0.001 § |
| Female | 461 (59.6%) | 352 (72.6%) | 22,987 (58.1%) |  |
| Male | 313 (40.4%) | 133 (27.4%) | 16,593 (41.9%) |  |
| **Age, years** |  |  |  | < 0.001 ¶ |
| Mean (SD) | 53.97 (15.54) | 58.56 (14.62) | 69.35 (14.57) |  |
| N-Miss | 1 | 0 | 0 |  |
| **Length of record, years** |  |  |  | 0.055 ¶ |
| Mean (SD) | 18.43 (11.21) | 19.41 (10.03) | 18.30 (10.18) |  |
| Range | 1.02 - 35.21 | 1.33 - 34.62 | 1.00 - 35.26 |  |
| **Insomnia** |  |  |  | < 0.001 § |
| Yes | 242 (33.8%) | 184 (41.3%) | 5601 (14.8%) |  |
| No | 473 (66.2%) | 262 (58.7%) | 32,302 (85.2%) |  |
| N-Miss<sup>b</sup> | 59 | 39 | 1677 |  |
| **Substance use disorder** |  |  |  | < 0.001 § |
| Yes | 230 (30.9%) | 122 (26.1%) | 845 (2.2%) |  |
| No | 514 (69.1%) | 346 (73.9%) | 38,361 (97.8%) |  |
| N-Miss<sup>‡</sup> | 30 | 17 | 374 |  |
| **Alcohol use disorder** |  |  |  | < 0.001 § |
| Yes | 205 (27.9%) | 99 (21.7%) | 1068 (2.7%) |  |
| No | 531 (72.1%) | 358 (78.3%) | 38,142 (97.3%) |  |
| N-Miss<sup>‡</sup> | 38 | 28 | 370 |  |
| **Tobacco use disorder** |  |  |  | < 0.001 § |
| Yes | 349 (47.5%) | 226 (48.4%) | 9138 (25.2%) |  |
| No | 385 (52.5%) | 241 (51.6%) | 27,161 (74.8%) |  |
| N-Miss<sup>‡</sup> | 40 | 18 | 3281 |  |

†Mayo Clinic Bipolar Disorder Biobank participants only included participants recruited at the Mayo Clinic
‡N-miss includes individuals with only 1 occurrence of the phecode
§Pearson's Chi-squared test
¶Linear Model ANOVA

The insomnia PRS was significantly associated with having an insomnia diagnosis in the EHR (i.e. at least 2 insomnia phecodes) in controls (OR=1.19, p=9.64e-33) but not in patients with BD (OR=1, p=0.95, Figure 1, Table S1). The interaction analysis confirmed that the effect was significantly greater in controls than cases (interaction p-value=0.006). In both cases and controls, the insomnia PRS was also found to be associated with diagnoses of SUDs (OR=1.34, p=8.39e-06 in cases and OR=1.11, p=2.47e-03 in controls), AUD (OR=1.22, p=4.35e-03 in cases OR=1.12, p=1.78e-04 in controls) and TUD (OR=1.2, p=2.38e-03 in cases and OR=1.11, p=3.6e-17 in controls). For the SUD phecode, a marginal difference was observed, such that the association with insomnia genetic liability was stronger in BD cases than controls (interaction p=0.024).

**Figure 1:**
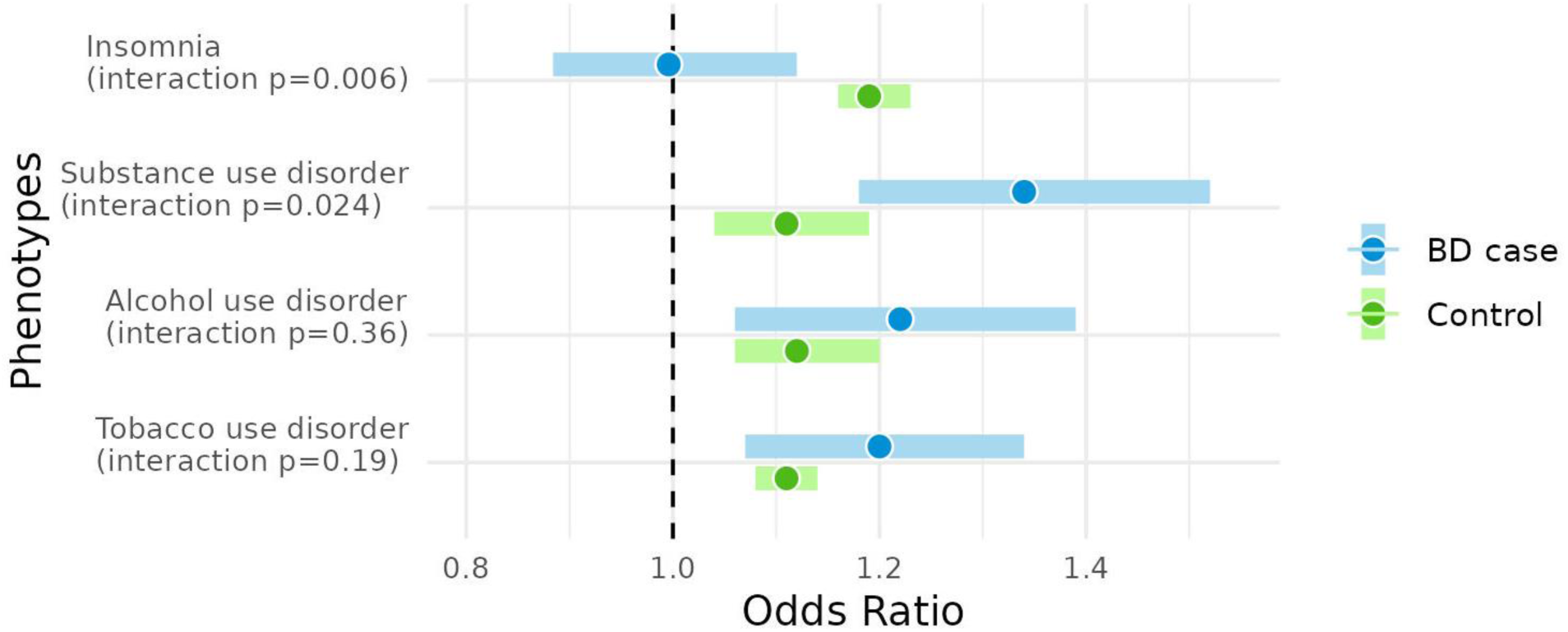
Forest plot of associations between insomnia PRS and EHR-derived phenotypes in BD cases and controls. Bars represent 95% confidence intervals of the odds ratio, which correspond to the odds of the phecode associated with a 1 SD increase in PRS.

We next investigated the association of insomnia PRS with SUDs in patients with BD (n=1789, Table 2) using the more detailed assessments of the BD Biobank. In this sample, insomnia PRS was associated with greater odds of BD-II subtype than BD-I (OR=1.2, 95% CI=1.09,1.33, p=3.84e-04). Higher insomnia PRS was also associated with increased odds of AUD (OR=1.19, p=4.26e-04), TUD (OR=1.21, p=1.25e-05) and CUD (OR=1.16, p=4.19e-03) as shown in Figure 2 and Table S2.

**Figure 2:**
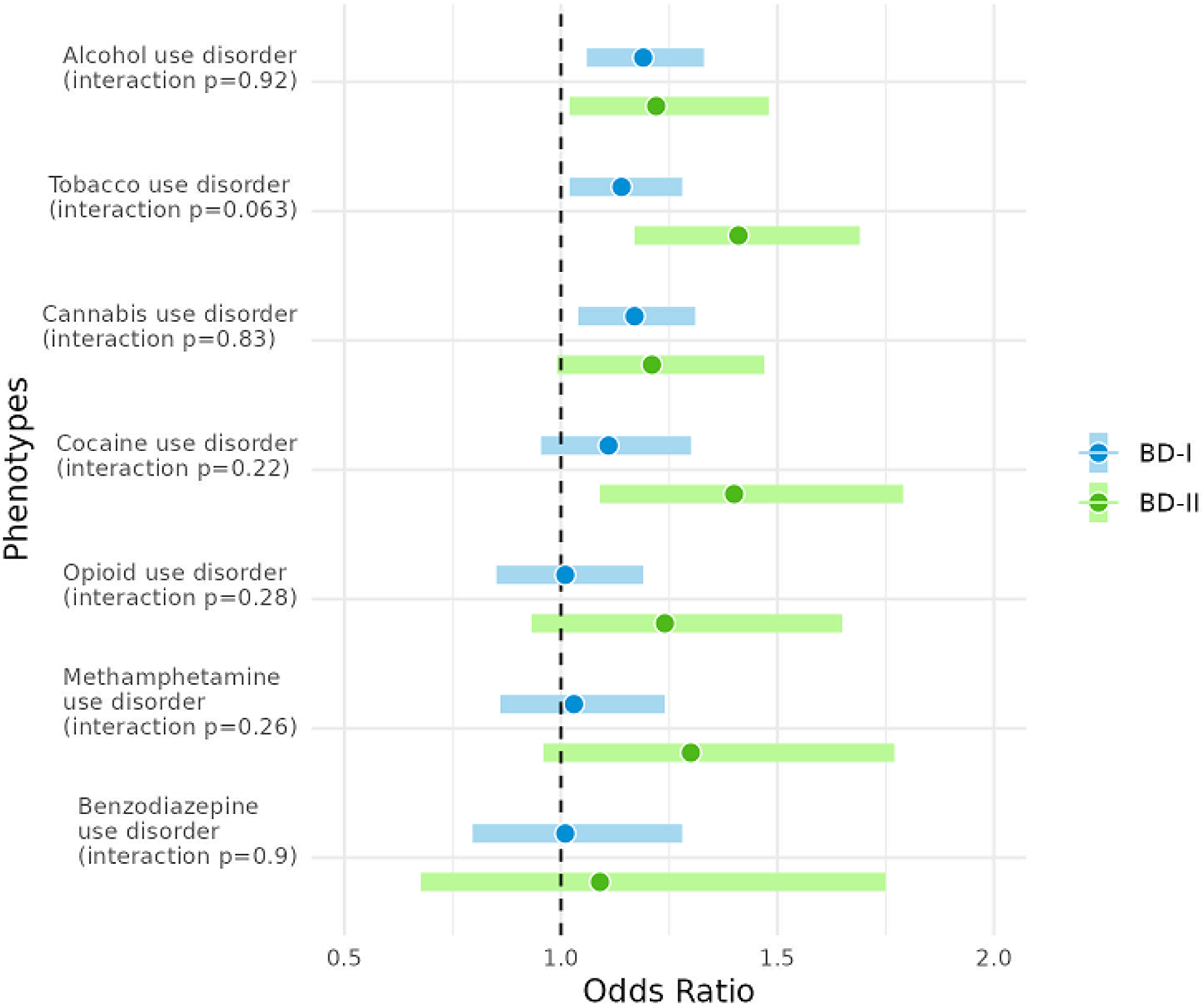
Forest plot of associations between insomnia PRS and SUD phenotypes in BD-I and BD-II cases. Bars represent 95% confidence intervals of the odds ratio, which correspond to the odds of the SUD phenotypes associated with a 1 SD increase in PRS.

**Table 2:** Demographics for the BD Biobank US sites used for analysis of questionnaire data.

|  | Overall (N=1789) |
| --- | --- |
| **Sex** |  |
| Female | 1092 (61.0%) |
| Male | 697 (39.0%) |
| **Age, yrs** |  |
| Mean (SD) | 43.17 (14.96) |
| Range | 18.00 - 80.00 |
| N-Miss | 8 |
| **Site** |  |
| Lindner Centre of Hope | 680 (38.0%) |
| Mayo Clinic Eau Claire | 2 (0.1%) |
| Mayo Clinic Florida | 22 (1.2%) |
| Mayo Clinic Rochester | 1020 (57.0%) |
| University of Minnesota | 65 (3.6%) |
| **Genotyping batch** |  |
| Batch 1 | 317 (17.7%) |
| Batch 2 | 471 (26.3%) |
| Batch 3 | 1001 (56.0%) |

In the BD-stratified analyses, (Figure 2, Table S2), significant associations were identified between insomnia PRS and AUD (OR=1.19, p=3.48e-03) in BD-I, and insomnia PRS and TUD (OR=1.41, p=2.83e-04) was found in BD-II. No significant interactions were identified, although the association of insomnia PRS with TUD was marginally stronger in BD-II than in BD-I (interaction p=0.063).

## Discussion

We investigated associations between insomnia genetic liability and SUDs in patients with BD compared to healthy controls using EHR-derived data, and further investigated similar associations in patients with BD using more detailed SUD data from the BD Biobank.

As expected, we found an association between insomnia PRS and the EHR-derived insomnia phenotype in the control population; however, we did not find an association in a sample of patients with BD. While this may partly reflect reduced power to detect an association in BD cases, an interaction analysis confirmed that the association of insomnia PRS with insomnia diagnosis was significantly weaker in BD, suggesting that the polygenic contribution to insomnia is greater in controls than cases. These results could indicate other risk factors contributing disproportionately to insomnia in the BD group, including other genetic risk factors and potential environmental factors. For example, substance use, including the use of psychotropic medications, is associated with insomnia symptoms ^10,38,39^. Moreover, a recent study found that BD cases with clinically significant insomnia experienced a greater number of early life stressors and impaired resilience than individuals with BD without insomnia ^40^, thereby suggesting the environment may have a greater contribution to the development of insomnia in BD.

Using EHR data, we demonstrated that insomnia PRS is also associated with SUD, AUD and TUD in BD and control subjects. This is in line with prior research that suggested a genetic predisposition for insomnia is associated with AUD ^21,41^. We were the first to incorporate insomnia, BD and SUDs in one analysis, thereby extending upon previous research, and demonstrating that insomnia PRS is associated with SUD, AUD and TUD not only in the general populations but also in patients with BD. More importantly, we observed an interaction between the insomnia PRS and BD case-control status for SUD, suggesting that the contribution of insomnia genetic liability to SUD is stronger in BD cases than controls. We also fit models that included interactions between our interaction terms of interest (insomnia PRS and BD) and other covariates in the model (sex and significant principal components) and obtained similar results (data not shown). Our findings suggest that the underlying biology of SUD risk may differ in individuals with BD and thus treatment strategy may need to be adapted for these individuals. We also note that the SUD phecode (316) encompasses several types of substances, including alcohol, cannabinoids, opioids, and stimulants, among others ^25^, and analyses in a larger sample are needed to determine which substance(s) is/are driving this interaction.

In the second part of the study, we leveraged SUD phenotype data from the BD Biobank to target questions we could not address using EHR data. First, we found that genetic liability to insomnia was associated with BD subtype, such that patients with BD-II had higher insomnia genetic liability than those with BD-I. These results are somewhat in accordance with a study that found that insomnia PRS was higher in BD-II than in controls but was not significantly different between BD-I and controls ^20^. A strong positive genetic correlation has also been reported between depression and insomnia ^16,42^, which further supports our findings given that depression has a stronger positive genetic correlation with BD-II than with BD-I ^17,43^.

Using SUD phenotypes from the BD Biobank questionnaire, we also confirmed the associations with AUD and TUD seen within BD cases for EHR-derived SUD phenotypes. Previous research has demonstrated shared common genetic liability between insomnia and nicotine dependence, insomnia and alcohol dependence as well as BD and smoking behaviors/nicotine dependence, which supports our findings ^16,44,45^. We also found insomnia PRS is associated with greater odds of CUD as well as marginally greater odds of cocaine use disorder. Given cocaine’s stimulating effects, it may be one of the substances driving the associations with the SUD phecode (316) in BD in our analyses using EHR-derived phenotypes. Given the novelty of this finding, additional research is warranted. Our exploratory analysis of differences in these associations across BD subtype was underpowered and did not detect any large differences. However, we did find marginal evidence suggesting that insomnia PRS may have a stronger association with TUD in BD-II than in BDI. While this result warrants replication, prior research has shown that smoking behaviors and nicotine dependence have a stronger positive genetic correlation with depression than schizophrenia ^44,45^. Given that depression is genetically correlated with BD ^17^, these results support the results from our exploratory analysis.

We acknowledge there are several limitations to this research. Analyses using phecodes rely on ICD codes, which may have potential inaccuracies between the true diagnosis and ICD code due to human error or diagnostic instability for psychiatric conditions ^46,47^. To overcome this limitation, we defined EHR-derived phenotypes based on at least two occurrences of a phecode in their EHR ^48,49^. It is important to recognize that SUD related ICD codes may be particularly underreported in EHR, possibly due to social stigma or underreporting to a clinician, which may have reduced power of the analyses. Additionally, our analyses on specific SUDs in the BD Biobank were based on self-reports of either a past or present SUD. Given the social stigma around some of the substances studied, SUDs may have been underreported, therefore leading to type II errors. Thus, the lack of insomnia PRS associations identified for OUD, methamphetamine use disorder, and benzodiazepine use disorder may be due to the lower prevalence of these conditions or their underreporting, rather than a true null effect ^50^. We also recognize that the smaller sample size of the BD-II group limited the power of the BD-type stratified analyses. Nevertheless, many of our results are supported by the depression literature, which is genetically highly correlated with BD-II ^17^. Finally, the results from this paper are based upon a PRS that was constructed using summary statistics from a GWAS of insomnia performed in the general population ^15^. Results may differ had we been able to use summary statistics from a GWAS of insomnia performed in a cohort of BD subjects.

## Conclusion

Findings from this study suggest that the effect of genetic predisposition to insomnia on SUD risk may be stronger in patients with BD, which may have important implications for clinical care for individuals with BD with comorbid SUD. Further work should focus on replicating and extending these results in large well-phenotyped cohorts.

## Data Availability

Data in the present study is available through application to each biobank and is unable to be shared otherwise.

## Acknowledgements

This work was supported by the Thomas and Elizabeth Grainger Fund in Bipolar Functional Genomics and Drug Development and National Institute of Mental Health R01 MH121924. The development of the Mayo Clinic Bipolar Disorder Biobank (BD Biobank) was supported by the Marriott Foundation, and the BD Biobank and the Mayo Clinic Biobank (MCB) were supported by the Mayo Clinic Center for Individualized Medicine.

We thank the MCB and BD Biobank research teams as well as the patient participants who volunteered their time to participate in these studies. We would also like to acknowledge Regeneron Genetics Center for providing genetic data for Mayo Clinic Biobank and some of the Mayo Bipolar Biobank participants.

## References

1. Bertrand L, Bourguignon C, Beaulieu S, Storch KF, Linnaranta O. Suicidal Ideation and Insomnia in Bipolar Disorders: Ideation suicidaire et insomnie dans les troubles bipolaires. Can J Psychiatry. Nov 2020;65(11):802–810. doi:10.1177/0706743720952226

2. Sublette EM, Carballo JJ, Moreno C, et al. Substance use disorders and suicide attempts in bipolar subtypes. J Psychiatr Res. Jan 2009;43(3):230–8. doi:10.1016/j.jpsychires.2008.05.001

3. Steinan MK, Scott J, Lagerberg TV, et al. Sleep problems in bipolar disorders: more than just insomnia. Acta Psychiatr Scand. May 2016;133(5):368–77. doi:10.1111/acps.12523

4. Palagini L, Cipollone G, Masci I, et al. Insomnia symptoms predict emotional dysregulation, impulsivity and suicidality in depressive bipolar II patients with mixed features. Compr Psychiatry. Feb 2019;89:46–51. doi:10.1016/j.comppsych.2018.12.009

5. Ritter PS, Hofler M, Wittchen HU, et al. Disturbed sleep as risk factor for the subsequent onset of bipolar disorder--Data from a 10-year prospective-longitudinal study among adolescents and young adults. J Psychiatr Res. Sep 2015;68:76–82. doi:10.1016/j.jpsychires.2015.06.005

6. Van Meter AR, Burke C, Youngstrom EA, Faedda GL, Correll CU. The Bipolar Prodrome: Meta-Analysis of Symptom Prevalence Prior to Initial or Recurrent Mood Episodes. J Am Acad Child Adolesc Psychiatry. Jul 2016;55(7):543–55. doi:10.1016/j.jaac.2016.04.017

7. Grunze H, Schaefer M, Scherk H, Born C, Preuss UW. Comorbid Bipolar and Alcohol Use Disorder-A Therapeutic Challenge. Front Psychiatry. 2021;12:660432. doi:10.3389/fpsyt.2021.660432

8. Hunt GE, Malhi GS, Cleary M, Lai HM, Sitharthan T. Prevalence of comorbid bipolar and substance use disorders in clinical settings, 1990-2015: Systematic review and meta-analysis. J Affect Disord. Dec 2016;206:331–349. doi:10.1016/j.jad.2016.07.011

9. Kenneson A, Funderburk JS, Maisto SA. Substance use disorders increase the odds of subsequent mood disorders. Drug Alcohol Depend. Dec 1 2013;133:338–343. doi:10.1016/j.drugalcdep.2013.06.011

10. Angarita GA, Emadi N, Hodges S, Morgan PT. Sleep abnormalities associated with alcohol, cannabis, cocaine, and opiate use: a comprehensive review. Addict Sci Clin Pract. Apr 26 2016;11(1):9. doi:10.1186/s13722-016-0056-7

11. Breslau N, Roth T, Rosenthal L, Andreski P. Sleep disturbance and psychiatric disorders: A longitudinal epidemiological study of young adults. Biological Psychiatry. 1996;39:411–418.

12. Hasler BP, Martin CS, Wood DS, Rosario B, Clark DB. A longitudinal study of insomnia and other sleep complaints in adolescents with and without alcohol use disorders. Alcohol Clin Exp Res. Aug 2014;38(8):2225–33. doi:10.1111/acer.12474

13. Chaudhary NS, Kampman KM, Kranzler HR, Grandner MA, Debbarma S, Chakravorty S. Insomnia in alcohol dependent subjects is associated with greater psychosocial problem severity. Addict Behav. Nov 2015;50:165–72. doi:10.1016/j.addbeh.2015.06.021

14. Kolla BP, Mansukhani MP, Biernacka J, Chakravorty S, Karpyak VM. Sleep disturbances in early alcohol recovery: Prevalence and associations with clinical characteristics and severity of alcohol consumption. Drug Alcohol Depend. Jan 1 2020;206:107655. doi:10.1016/j.drugalcdep.2019.107655

15. Lane JM, Jones SE, Dashti HS, et al. Biological and clinical insights from genetics of insomnia symptoms. Nat Genet. Mar 2019;51(3):387–393. doi:10.1038/s41588-019-0361-7

16. Watanabe K, Jansen PR, Savage JE, et al. Genome-wide meta-analysis of insomnia prioritizes genes associated with metabolic and psychiatric pathways. Nat Genet. Aug 2022;54(8):1125–1132. doi:10.1038/s41588-022-01124-w

17. Mullins N, Forstner AJ, O’Connell KS, et al. Genome-wide association study of more than 40,000 bipolar disorder cases provides new insights into the underlying biology. Nat Genet. Jun 2021;53:817–829. doi:10.1038/s41588-021-00857-4

18. Zhou H, Rentsch CT, Cheng Z, et al. Association of OPRM1 Functional Coding Variant With Opioid Use Disorder: A Genome-Wide Association Study. JAMA Psychiatry. Oct 1 2020;77(10):1072–1080. doi:10.1001/jamapsychiatry.2020.1206

19. Kranzler HR, Zhou H, Kember RL, et al. Genome-wide association study of alcohol consumption and use disorder in 274,424 individuals from multiple populations. Nature Communications. 2019;10(1)doi:10.1038/s41467-019-09480-8

20. Lewis KJS, Richards A, Karlsson R, et al. Comparison of Genetic Liability for Sleep Traits Among Individuals With Bipolar Disorder I or II and Control Participants. JAMA Psychiatry. Mar 1 2020;77(3):303–310. doi:10.1001/jamapsychiatry.2019.4079

21. Chakravorty S, Kember RL, Mazzotti DR, et al. The relationship between alcohol- and sleep-related traits: Results from polygenic risk score and Mendelian randomization analyses. Drug Alcohol Depend. Oct 1 2023;251:110912. doi:10.1016/j.drugalcdep.2023.110912

22. Olson JE, Ryu E, Johnson KJ, et al. The Mayo Clinic Biobank: a building block for individualized medicine. Mayo Clin Proc. Sep 2013;88(9):952–62. doi:10.1016/j.mayocp.2013.06.006

23. Frye MA, McElroy SL, Fuentes M, et al. Development of a bipolar disorder biobank: differential phenotyping for subsequent biomarker analyses. Int J Bipolar Disord. Dec 2015;3:14. doi:10.1186/s40345-015-0030-4

24. First MB, Gibbon M. The Structured Clinical Interview for DSM-IV Axis I Disorders (SCID-I) and the Structured Clinical Interview for DSM-IV Axis II Disorders (SCID-II). Comprehensive handbook of psychological assessment*, Vol* 2: Personality assessment. John Wiley & Sons, Inc.; 2004:134–143.

25. Wu P, Gifford A, Meng X, et al. Mapping ICD-10 and ICD-10-CM Codes to Phecodes: Workflow Development and Initial Evaluation. Original Paper. JMIR Med Inform. 2019;7(4):e14325. doi:10.2196/14325

26. Denny JC, Ritchie MD, Basford MA, et al. PheWAS: demonstrating the feasibility of a phenome-wide scan to discover gene-disease associations. Bioinformatics. May 1 2010;26(9):1205–10. doi:10.1093/bioinformatics/btq126

27. Denny JC, Bastarache L, Ritchie MD, et al. Systematic comparison of phenome-wide association study of electronic medical record data and genome-wide association study data. Nat Biotechnol. Dec 2013;31(12):1102–10. doi:10.1038/nbt.2749

28. Coombes BJ, Landi I, Choi KW, et al. The genetic contribution to the comorbidity of depression and anxiety: a multi-site electronic health records study of almost 178 000 people. Psychol Med. Nov 2023;53(15):7368–7374. doi:10.1017/S0033291723000983

29. Das S, Forer L, Schonherr S, et al. Next-generation genotype imputation service and methods. Nat Genet. Oct 2016;48(10):1284–1287. doi:10.1038/ng.3656

30. Loh PR, Danecek P, Palamara PF, et al. Reference-based phasing using the Haplotype Reference Consortium panel. Nat Genet. Nov 2016;48(11):1443–1448. doi:10.1038/ng.3679

31. Fuchsberger C, Abecasis GR, Hinds DA. minimac2: faster genotype imputation. Bioinformatics. Mar 1 2015;31(5):782–4. doi:10.1093/bioinformatics/btu704

32. Taliun D, Harris DN, Kessler MD, et al. Sequencing of 53,831 diverse genomes from the NHLBI TOPMed Program. Nature. Feb 2021;590(7845):290–299. doi:10.1038/s41586-021-03205-y

33. Manichaikul A, Mychaleckyj JC, Rich SS, Daly K, Sale M, Chen WM. Robust relationship inference in genome-wide association studies. Bioinformatics. Nov 15 2010;26(22):2867–73. doi:10.1093/bioinformatics/btq559

34. Alexander DH, Lange K. Enhancements to the ADMIXTURE algorithm for individual ancestry estimation. BMC Bioinformatics. 2011;(12):246.

35. 1000 Genomes Project Consortium, Auton A, Brooks LD, et al. A global reference for human genetic variation. Nature. Oct 1 2015;526(7571):68–74. doi:10.1038/nature15393

36. Abraham G, Qiu Y, Inouye M. FlashPCA2: principal component analysis of Biobank-scale genotype datasets. Bioinformatics. Sep 1 2017;33(17):2776–2778. doi:10.1093/bioinformatics/btx299

37. Prive F, Albinana C, Arbel J, Pasaniuc B, Vilhjalmsson BJ. Inferring disease architecture and predictive ability with LDpred2-auto. Am J Hum Genet. 2023;110(12):2042–2055. doi:10.1016/j.ajhg.2023.10.010

38. Merrill RM, Ashton MK, Angell E. Sleep disorders related to index and comorbid mental disorders and psychotropic drugs. Ann Gen Psychiatry. May 27 2023;22(1):23. doi:10.1186/s12991-023-00452-3

39. Glastad SH, Aminoff SR, Hagen R, et al. Nicotine use and non-pathological alcohol use and their relationship to affective symptoms and sleep disturbances in bipolar disorder. J Affect Disord. Apr 14 2023;327:236–243. doi:10.1016/j.jad.2023.02.003

40. Palagini L, Miniati M, Marazziti D, et al. Insomnia symptoms are associated with impaired resilience in bipolar disorder: Potential links with early life stressors may affect mood features and suicidal risk. J Affect Disord. 2022;299:596–603. doi:10.1016/j.jad.2021.12.042

41. Reginsson GW, Ingason A, Euesden J, et al. Polygenic risk scores for schizophrenia and bipolar disorder associate with addiction. Addict Biol. Jan 2018;23(1):485–492. doi:10.1111/adb.12496

42. Howard DM, Adams MJ, Clarke T-K, et al. Genome-wide meta-analysis of depression identifies 102 independent variants and highlights the importance of the prefrontal brain regions. Nature Neuroscience. 2019;22(3):343–352. doi:10.1038/s41593-018-0326-7

43. Song J, Bergen SE, Kuja-Halkola R, Larsson H, Landen M, Lichtenstein P. Bipolar disorder and its relation to major psychiatric disorders: a family-based study in the Swedish population. Bipolar Disorders. Mar 2015;17:184–193. doi:10.1111/bdi.12242

44. Barkhuizen W, Dudbridge F, Ronald A. Genetic overlap and causal associations between smoking behaviours and mental health. Sci Rep. Jul 21 2021;11(1):14871. doi:10.1038/s41598-021-93962-7

45. Abdellaoui A, Smit DJA, van den Brink W, Denys D, Verweij KJH. Genomic relationships across psychiatric disorders including substance use disorders. Drug and Alcohol Dependence. 2021;220doi:10.1016/j.drugalcdep.2021.108535

46. Baca-Garcia E, Perez-Rodriguez MM, Basurte-Villamor I, et al. Diagnostic stability of psychiatric disorders in clinical practice. Br J Psychiatry. Mar 2007;190:210–216. doi:10.1192/bjp.bp.106.024026

47. Bastarache L. Using phecodes for research with the electronic health record: From PheWAS to PheRS. Annu Rev Biomed Data Sci. Jul 20 2021;4:1–19. doi:10.1146/annurev-biodatasci-122320-112352

48. Sanchez-Roige S, Fontanillas P, Jennings MV, et al. Genome-wide association study of problematic opioid prescription use in 132,113 23andMe research participants of European ancestry. Mol Psychiatry. Nov 2021;26(11):6209–6217. doi:10.1038/s41380-021-01335-3

49. Zheutlin AB, Dennis J, Linnér RK, et al. Penetrance and pleiotropy of polygenic risk scores for schizophrenia in 90,000 patients across three healthcare systems. 2019;doi:10.1101/421164

50. Steinhoff A, Shanahan L, Bechtiger L, et al. When Substance Use Is Underreported: Comparing Self-Reports and Hair Toxicology in an Urban Cohort of Young Adults. J Am Acad Child Adolesc Psychiatry. Jul 2023;62(7):791–804. doi:10.1016/j.jaac.2022.11.011

