## Supplemental Figure 1 for "Genetic liability to insomnia and substance use disorders in patients with bipolar disorder"

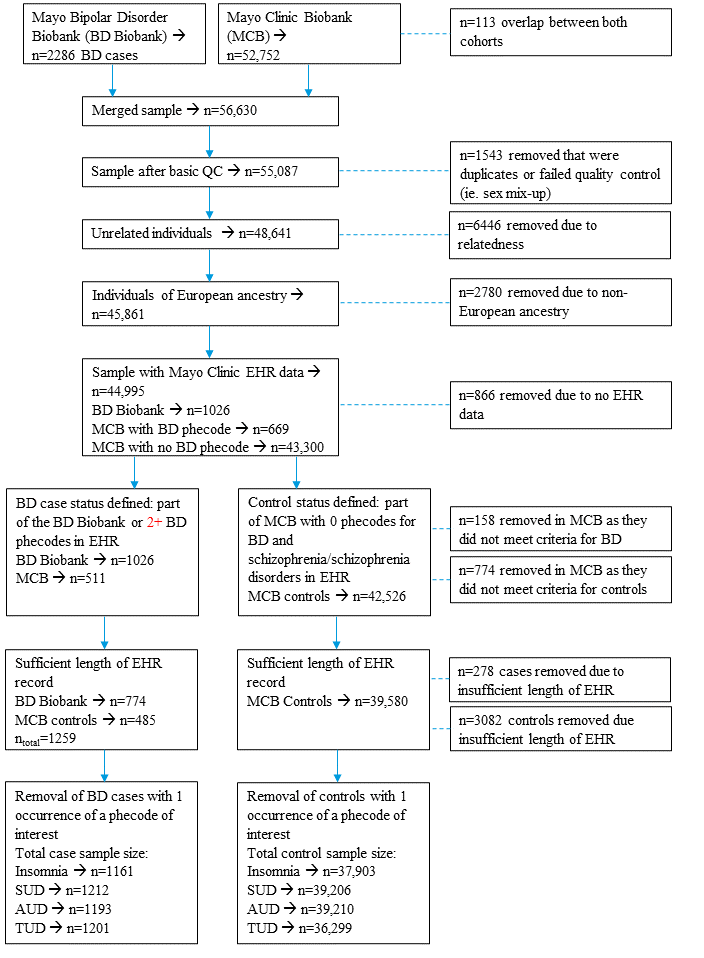

Figure S1: Overview of sample refinement for association analyses between insomnia PRS and EHR-derived phenotypes
